# Evidence for reduced somatic T-cell receptor sequence diversity profiles among Midwestern Amish in an aging cohort study

**DOI:** 10.64898/2026.01.25.26344754

**Authors:** Lauren A. Cruz, Shiying Liu, Kristy L. Miskimen, Jessica N. Cooke Bailey, Tyler Kinzy, Yeunjoo E. Song, Renee A. Laux, Penelope Miron, Paula K. Ogrocki, Alan J. Lerner, Audrey Lynn, Sarada L. Fuzzell, Sherri D. Hochstetler, Ioanna Konidari, Jacob L. McCauley, William K. Scott, Margaret A. Pericak-Vance, Jonathan L. Haines, Dana C. Crawford

**Affiliations:** Department of Population and Quantitative Health Sciences, Cleveland Institute for Computational Biology, Case Western Reserve University School of Medicine, Cleveland, Ohio; East Carolina University, Greenville, NC; Department of Neurology, University Hospitals Cleveland Medical Center, Cleveland, Ohio; The Dr. John T. Macdonald Foundation Department of Human Genetics, John P. Hussman Institute for Human Genomics, Miller School of Medicine, University of Miami, Miami, Florida

**Keywords:** Dementia, Adaptive Immunity, Human Leukocyte Antigen, Genetics

## Abstract

Late-onset Alzheimer disease (LOAD), the most common form of dementia among older adults, is a neurodegenerative disease characterized by brain amyloid-β (Aβ) plaque deposition and neurofibrillary tangles. The causes of LOAD are not known but several recent lines of evidence implicate the adaptive immune system. Here, we sought to characterize somatic T-cell receptor (TCR) sequence diversity profiles and class I and II human leukocyte antigen (HLA) alleles from DNA extracted from peripheral tissues from Midwestern Amish participating in longitudinal studies of aging. We immunosequenced the TCR beta chain from genomic DNA of 72 Midwestern Amish, including participants with clinically diagnosed LOAD (n=6), mild cognitive impairment (MCI; n=16), cognitive impairment but not AD (CINAD; n=3), and 35 cognitively unaffected. TCR sequence diversity by cognitive status was examined using a variety of metrics, and tests of association were performed between cognitive status and HLA alleles. For a subset of participants, plasma biomarkers for LOAD pathogenesis were available to evaluate TCR sequence diversity by cognitive status. TCR sequence diversity measured as Simpson’s clonality was lower among LOAD+MCI compared with non-LOAD, but these differences were not independent of age. Relatively few clonotypes (exact nucleotide sequences) were shared across participants; of those few shared include the Epstein Barr virus associated clonotype. HLA-A*03:01 and several HLA-DRB1 alleles were under-represented among LOAD+MCI participants compared with cognitively unaffected participants, but these associations were no longer significant in adjusted analyses. Among LOAD+MCI participants with plasma biomarkers, increased p-tau181 was associated decreased TCR sequence diversity, and the association was independent of age. In this limited Midwestern Amish sample, the observed TCR diversity associations are consistent with the involvement of the adaptive immune system in LOAD.

## Introduction

Late-onset Alzheimer disease (LOAD) is a progressive neurodegenerative disease and the most common cause of dementia. LOAD is the sixth leading cause of mortality in the United States (US) ("2024 Alzheimer’s disease facts and figures," 2024) and the 5th leading causes of death among those ≥ 65 years of age (Tejada-Vera, 2013). Approximately 6.7 million people residing in the US ≥65 years of age are currently living with LOAD ("2024 Alzheimer’s disease facts and figures," 2024). The estimated annual cost of dementia in 2025 is $781 billion USD (Team, 2025), and the cost is expected to rise to the trillions by 2060 (Nandi et al., 2024). No cures exist for LOAD, and it is not yet certain if recent FDA-approved therapies will be substantially effective in slowing clinical progression among those affected (Beveridge et al., 2024).

LOAD is a major neurodegenerative disorder, and its prevalence varies by US population (Steenland et al., 2016; Tang et al., 2001). The Amish, groups of German and Swiss Anabaptists who immigrated to the US in the 1700’s and 1800’s, have lower rates of LOAD compared with European-descent residents drawn from the general US population (D’Aoust et al., 2015; Johnson et al., 1997). The lower frequency of *APOE e4*, the major known genetic risk factor for LOAD, does not account for the lower LOAD prevalence among the Amish (Pericak-Vance et al., 1996). Social determinants of health such as healthcare and education access and quality (Ramos et al., 2021), also do not explain the differences in LOAD prevalence. As a uniformly uninsured rural group with on average an eighth-grade education and limited access to consistent quality medical care (Ramos et al., 2021), the Amish provide a unique opportunity to identify trans-population and population-specific risk and protective factors associated with LOAD among aging populations.

LOAD is a complex disease influenced by a combination of genetic and environmental risk factors (Scheltens et al., 2016). Modifiable factors include education (Ramos et al. 2021), smoking (Norton et al., 2014), and obesity (Kivipelto et al., 2005). Known non-modifiable risk factors include female sex, increased age, and LOAD-associated genetic risk alleles (Bellenguez et al., 2022), most notably *APOE e4* (Corder et al., 1995). *APOE* normally encodes a product that regulates cholesterol in the brain, and it is known to bind to the Aβ peptide (Holtzman et al., 2012).

In addition to known risk factors, both the innate and adaptive immune systems have been implicated in LOAD risk (Bradshaw et al., 2013; Guerreiro et al., 2013; Jevtic et al., 2017; Labzin et al., 2018; Li et al., 2021; Marsh et al., 2016; Sims et al., 2017). Further, LOAD may be a T-cell mediated disease (Dressman & Elyaman, 2022) that can be characterized by T-cell receptor (TCR) sequences or repertoires. The strongest evidence for TCR repertoire signatures in LOAD is based on characterization of sequences from brain tissue (e.g, (Chen et al., 2023). While these samples are representative of the affected tissue, their routine collection from living participants or patients is not feasible. Peripheral tissues such as blood and saliva are easier to collect and are an attractive target for population-scale studies and clinical screening. While LOAD-associated immune profiles have been detected in blood (Shigemizu et al., 2022), additional data are needed in different populations drawn from different genetic backgrounds and environmental contexts. In this study, we sought to understand the role of adaptive immunity in the LOAD pathophysiology in an older Midwestern Amish rural cohort, with the primary objectives of the characterization of TCR repertoire profile using DNA from peripheral tissues and the examination of the Major Histocompatibility Complex (MHC) genomic region of the adaptive immune system in both affected and unaffected individuals.

## Methods

Samples for immunosequencing were primarily selected to represent LOAD cases (“affected”) and non-cases (“unaffected”) for immunosequencing from the larger Collaborative Amish & Aging Memory Project (CAAMP), a longitudinal study of cognitively normal but at-risk Amish adults ≥75 years of age or existing participants’ siblings ≥65 years of age living in Holmes County, Ohio and Adams, Elkhart, and LaGrange Counties, Indiana. Present communities in Holmes (Ohio) and Elkhart/LaGrange (Indiana) counties are primarily descendants from the first immigration wave from the German Palatinate between 1727 and 1770, while the Amish of Adams (Ohio) county are principally descendants from the second immigration from Switzerland between 1815 and 1860 (Agarwala et al., 2001; Hostetler, 1993; Jackson et al., 1968). Informed consent was obtained from Amish participants, and the study and its protocols were approved by an appropriate Internal Review Board (University Hospitals CR00007198).

### Study Population

A subset of CAAMP participants were selected for the present study based on consensus LOAD-related phenotype at the time of sequencing, the availability of sufficient genomic DNA, and availability of genome-wide array data. A total of 72 participants were immunosequenced (Table 1). Biomarker data, including Aβ40, Aβ42, total tau (t-tau), and phosphorylated tau protein at epitope 181 (p-tau181) were available for 61 participants obtained from frozen plasma samples that passed quality control procedures, excluding samples with intra-assay coefficient of variation > 20% and outliers evaluated based on median absolute aviation, as described by Wang et al (P. Wang et al., 2025).

**Table 1.** Study population characteristics. Age at sampling represents age of participant at the time of blood draw or saliva collection, while age at last exam represents age at most recent cognitive assessment. Affected (LOAD+MCI) versus unaffected sex, mean ages, and *APOE e4* status were compared using Fisher’s exact and two-tailed t-tests assuming unequal variances, where appropriate. Abbreviations: Late-onset Alzheimer disease (LOAD), cognitively impaired but not Alzheimer disease (CINAD), mild cognitive impairment (MCI), p-value (p), standard deviation (SD)

|  | **Total**<br>**(n=72)** | **Unaffected**<br>**(n=35)** | **Unclear**<br>**(n=12)** | **CINAD**<br>**(n=3)** | **MCI**<br>**(n=16)** | **LOAD**<br>**(n=6)** | **p** |
| --- | --- | --- | --- | --- | --- | --- | --- |
| Mean age at sampling ± SD | 79.9 ± 4.9 | 78.4 ± 4.4 | 79.7 ± 5.3 | 83.67 ± 4.0 | 81.8 ± 3.9 | 81.7 ± 7.7 | 0.01 |
| Mean age at last exam ± SD | 82.2 ± 4.0 | 81.5 ± 4.0 | 81.1 ± 3.7 | 85.0 ± 2.65 | 83.6 ± 4.0 | 84.7 ± 4.1 | 0.06 |
| % Female | 55.6 | 65.7 | 33.3 | 66.7 | 50.0 | 50.0 | 0.28 |
| Mean years education | 8.01 ± 0.2 | 8.06 ± 0.2 | 8.0 ± 0 | 8.0 ± 0 | 7.94 ± 0.3 | 8.0 ± 0 | 0.10 |
| % *APOE e2* | 3.5 | 5.7 | 4.2 | - | - | - |  |
| % *APOE e3* | 87.5 | 82.9 | 83.3 | 100 | 96.9 | 91.7 |  |
| % *APOE e4* | 9.0 | 11.4 | 12.5 | - | 3.1 | 8.3 | 0.31 |

### Phenotyping

CAAMP participants undergo cognitive screening using a battery of tests including the modified mini-mental state (3MS) education-adjusted examination score (Teng & Chui, 1987), the AD8 Dementia Screening Interviews (Galvin et al., 2007), the CERAD word list learning test (Welsh et al., 1994), and the Trailmaking test (Tombaugh, 2004). Other data including medical (via a checklist to document self-reported major chronic illnesses), family and health history, self-reported behavior and lifestyle (e.g., smoking), and functional assessments are collected from participants and their families.

A per-participant consensus phenotype was determined by a clinical adjudication board (CAB). The CAB reviews all clinical data to classify participants as cognitively normal or “unaffected” based on established thresholds for the administered tests (Teng & Chui, 1987; Tombaugh, 2004). For those participants who were not classified as “unaffected” using the established thresholds, additional evaluations were conducted including a full neurocognitive evaluation using a multi-test CERAD battery (Welsh et al., 1991; Welsh et al., 1994). From these data, the CAB generates consensus diagnoses for participants with memory and cognitive problems (Albert et al., 2011; McKhann et al., 2011; Osterman et al., 2023; Ramos et al., 2021). CAAMP participants receive follow-up exams every two years. All 72 participants in the present study have had one exam, and while most have had two exams (81.9%), few have had three exams (15.3%). Based on the most recent CAB consensus phenotype, the present study includes participants deemed cognitively unimpaired (unaffected, n=35), cognitively impaired but not AD (CINAD, n=3), mild cognitive impairment (MCI, n=16), LOAD (n=6), and unclear (n=12) (Table 1).

### Immunosequencing and genotyping

All biospecimens were collected from consented participants prior to the emergence of COVID-19. DNA from 72 Amish participants was extracted from peripheral tissues, including whole blood (99%) or saliva (1%), using Qiagen’s QIAsymphony. TCR beta chain CDR3 regions were amplified and barcoded in a two-step multiplex PCR using Adaptive Biotechnologies’ immunoSEQ kit (Carlson et al., 2013). Amplicons were sequenced using Illumina’s NextSeq. As recommended by Adaptive Biotechnologies, each DNA sample was sequenced using six replicates. Amplicon sequences were submitted to Adaptive Biotechnologies ANALYZER bioinformatics pipeline for quality control, alignment, and downstream analyses as described below. Genome-wide genetic data are available for participants, which include a mix of array-based data from the Illumina Global Screening Array with AD-associated custom content and Multi-ethnic global array (MEGA^EX^) plus 3K. Quality control was previously described (Osterman et al., 2022) and includes standard parameters (genotype call rates, batch effects, heterozygosity, sex-checks, and Mendelian error checks (Turner et al., 2011)). Given that multiple arrays were used to generate the data, standardized allele calling and strand alignment with external databases was also performed. *APOE* was directly genotyped using TaqMan (Thermo Fisher Scientific) (Cummings et al., 2012).

### Statistical methods

Descriptive statistics include basic Fisher’s exact, Welch’s t-test, and logistic regression (R-4.4.1, R Studio version: 2024.04.2+764), where indicated. Basic immunosequencing metrics are provided by the Adaptive Biotechnologies ANALYZER, including the total templates, productive templates, and productive rearrangements by DNA sample. TCR sequence diversity is described using a variety of metrics, including productive Simpson’s clonality, Pielou evenness, and Morisita-Horn Index, among other diversity metrics commonly used in immunology and ecology (Chiffelle et al., 2020). Productive Simpson’s clonality index is a general measure of clonal evenness, also known as relative abundance. Values range from 0 to 1, where 0 represents a repertoire of completely even sequences (polyclonality), and 1 represents a single dominant clone or a few clones (monoclonality and oligoclonality, respectively). Lower values indicate more diversity, whereas higher values indicate reduced diversity. To study the association between the plasma biomarkers and TCR diversity, we performed linear regression stratified for LOAD disease status both with and without adjustment for age and sex. P-values were not corrected for multiple testing.

We employed the Michigan Imputation Server 2 (MIS) (Das et al., 2016) to impute human leukocyte antigen (HLA) class I and class II haplotypes for the genotyped Amish participants. Prior to imputation, we prepared the data using recommended pre-imputation checks against the MIS reference panel, and the data after modifications were then phased using Eagle v2.4. Imputation was performed using minimac4 leveraging the four-digit multiethnic HLA reference panel v2, which has >20,000 samples, providing 517 HLA alleles along with critical information for HLA amino acids and HLA SNPs (Luo et al., 2021). Post-imputation quality control was performed using the criteria to retain HLA alleles with an imputation quality score (R2) > 0.7 and minor allele frequency (MAF) >0.01, resulting in a total of 161 HLA alleles including both 2 and 4 digits. The subsequent HLA association tests and visualizations were accomplished leveraging methods adapted from HLA-Typing At Protein for Association Studies (HLA-TAPAS) (Luo et al., 2021), where we evaluated the associations both unadjusted and adjusted for age and sex.

## Results

We characterized somatic TCR sequence diversity profiles in clinically adjudicated LOAD (n=6), mild cognitive impairment (MCI; n=16), cognitively impaired but not Alzheimer disease (CINAD; n=3), unclear (n=12), and unaffected (n=35) Amish participants residing in Ohio and Indiana. As shown in Table 1, more than half of the 72 participants were female (55.6%), and MCI and LOAD (mean 81.8 and 81.7 years, respectively) participants were on average older at sampling compared with unaffected participants (78.4 years; p=0.01). Unaffected participants on average had more years of education (8.06) compared with affected (LOAD+MCI; 7.95 years ± 0.21 standard deviation) participants, but this difference was not statistically significant (p=0.10). *APOE 3* was common across all participants (87.5%), and the proportion carrying *APOE 4* did not differ between unaffected and affected (LOAD+MCI) participants (Table 1).

Sequencing was performed using Adaptive Biotechnologies immunoSEQ targeting the beta chain, resulting in >515,000 productive templates and >250,000 productive rearrangements (Table 2 and Supplementary Table 1). TCR sequence diversity, characterized by productive Simpson’s clonality, was lower among LOAD+MCI (0.12) compared with non-LOAD (0.09) participants, and this difference (t-test, p=0.04) remained after removal of one LOAD+MCI participant whose data were based on a saliva sample. However, the difference in average productive Simpson’s clonality between LOAD+MCI and non-LOAD participants did not remain significant after adjusting for age at sampling (p=0.148) or age at last exam (p=0.089). TCR diversity among participants with unclear diagnosis was intermediate (productive Simpson’s clonality=0.10).

**Table 2.** Summary of sequence-level metrics, by consensus phenotype status. All metrics save for Shannon diversity were provided by Adaptive Biotechnologies ANALYZER bioinformatic pipeline. Shown are means (standard deviations). Unaffected participants were compared with affected participants (LOAD+MCI) using Welch’s t-test. Abbreviations: Late-onset Alzheimer disease (LOAD), mild cognitive impairment (MCI), p-value (p).

|  | Unaffected<br>(n=35) | Affected<br>(n=22) |  | p |
| --- | --- | --- | --- | --- |
|  |  | MCI<br>(n=16) | LOAD<br>(n=6) |  |
| Total templates | 85,945.86<br>(33,212.27) | 87,733.94<br>(39,440.47) | 99,155.00<br>(32,021.43) | 0.62 |
| Productive templates | 68,218.6<br>(26,157.79) | 70,554.06<br>(31,014.97) | 78,092.17<br>(28,729.09) | 0.57 |
| Fraction productive templates | 0.79<br>(0.07) | 0.79<br>(0.09) | 0.78<br>(0.05) | 0.98 |
| Total rearrangements | 46,939.43<br>(19,079.45) | 39,836.50<br>(23,903.85) | 40,825.33<br>(14,022.59) | 0.23 |
| Productive rearrangements | 37,694.89<br>(15,495.43) | 32,021.81<br>(18,810.90) | 32,482.33<br>(11,542.84) | 0.22 |
| Maximum productive frequency | 0.06<br>(0.05) | 0.09<br>(0.07) | 0.15<br>(0.08) | 0.08 |
| Productive Simpson's clonality | 0.08<br>(0.06) | 0.12<br>(0.07) | 0.11<br>(0.08) | 0.04 |
| Pielou evenness | 0.84<br>(0.09) | 0.80<br>(0.12) | 0.75<br>(0.14) | 0.03 |

To examine the relationship between TCR diversity and plasma biomarkers relevant to amyloid and tau pathologies, we stratified by affected status and tested for unadjusted and adjusted associations with productive Simpson’s clonality. Among affected LOAD+MCI participants, productive Simpson’s clonality was associated with p-tau181 levels (p=0.047; Table 3). Despite the limited sample size (n=16), this association remained significant after adjusting for age at last exam and sex (p=0.011; Table 3). We also observed a significant association adjusted for age at last exam and sex between p-tau181 and productive Simpson’s clonality in non-LOAD participants (p=0.034; Table 3); however, the effect is in the opposite direction compared to the LOAD+MCI group. Significant associations were also observed between productive Simpson’s clonality and both Aβ42 levels and the Aβ42/Aβ40 ratio in adjusted analyses among LOAD+MCI participants but not among unaffected participants (Table 3).

**Table 3.** Productive Simpson’s clonality associations with biomarkers. Statistical tests were performed to investigate the association between productive Simpson’s clonality and a series of biomarker measurements, within affected (LOAD+MCI) and unaffected participants with biomarker measurements available. Both unadjusted **(a)** and adjusted **(b)** (for age at last exam and sex) analyses are presented. P-values shown are uncorrected for multiple testing.

**(a)**
|  | LOAD+MCI |  |  | Unaffected |  |  |
| --- | --- | --- | --- | --- | --- | --- |
|  | n | beta | p value | n | beta | p value |
| Aβ40 (pg/ml) | 16 | 119.87 | 0.694 | 33 | 108.07 | 0.733 |
| Aβ42 (pg/ml) | 13 | -19.18 | 0.191 | 32 | 6.56 | 0.688 |
| Aβ42/ Aβ40 | 13 | -0.07 | 0.073 | 32 | -0.06 | 0.627 |
| tau (pg/ml) | 13 | 2.54 | 0.589 | 25 | 3.52 | 0.283 |
| p-tau181 (pg/ml) | 16 | 8.63 | 0.047 | 33 | -4.04 | 0.068 |

**(b)**
|  | LOAD+MCI |  |  | Unaffected |  |  |
| --- | --- | --- | --- | --- | --- | --- |
|  | n | beta | p value | n | beta | p value |
| Aβ40 (pg/ml) | 16 | 88.23 | 0.785 | 33 | -19.08 | 0.953 |
| Aβ42 (pg/ml) | 13 | -31.73 | 0.037 | 32 | 1.08 | 0.950 |
| Aβ42/ Aβ40 | 13 | -0.09 | 0.026 | 32 | -0.05 | 0.648 |
| tau (pg/ml) | 13 | 2.16 | 0.659 | 25 | 3.14 | 0.417 |
| p-tau181 (pg/ml) | 16 | 10.53 | 0.011 | 33 | -4.88 | 0.034 |

We also examined the frequency of recurrent sequences for evidence of recent clonal expansion. Overall, affected (LOAD+MCI) participants had more dominant clones (defined here as productive rearrangements greater ≥10% in frequency) compared with unaffected participants (50% versus 25.7%, respectively). This pattern is suggestive of increased clonal expansion among LOAD+MCI compared with unaffected participants (Fisher’s exact, p=0.088) and is supported by the lower Pielou evenness when compared with unaffected participants (p=0.03; Table 1). Unclear participants (33.3%) had more dominant clones than unaffected participants but fewer than affected (LOAD+MCI). Relatively few clonotypes (exact nucleotide sequences) were shared across participants. Pairwise comparisons of sequence overlap between individuals revealed few sequence overlaps indicating greater sample uniqueness compared between individuals using the Morisita-Horn index (Figure 1). Among the few shared clonotypes identified was the Epstein Barr virus associated clonotype (CASSLGQAYEQYF), with at least one count detected in 36% and 45% of the LOAD+MCI and cognitively unaffected participants, respectively.

**Figure 1.**
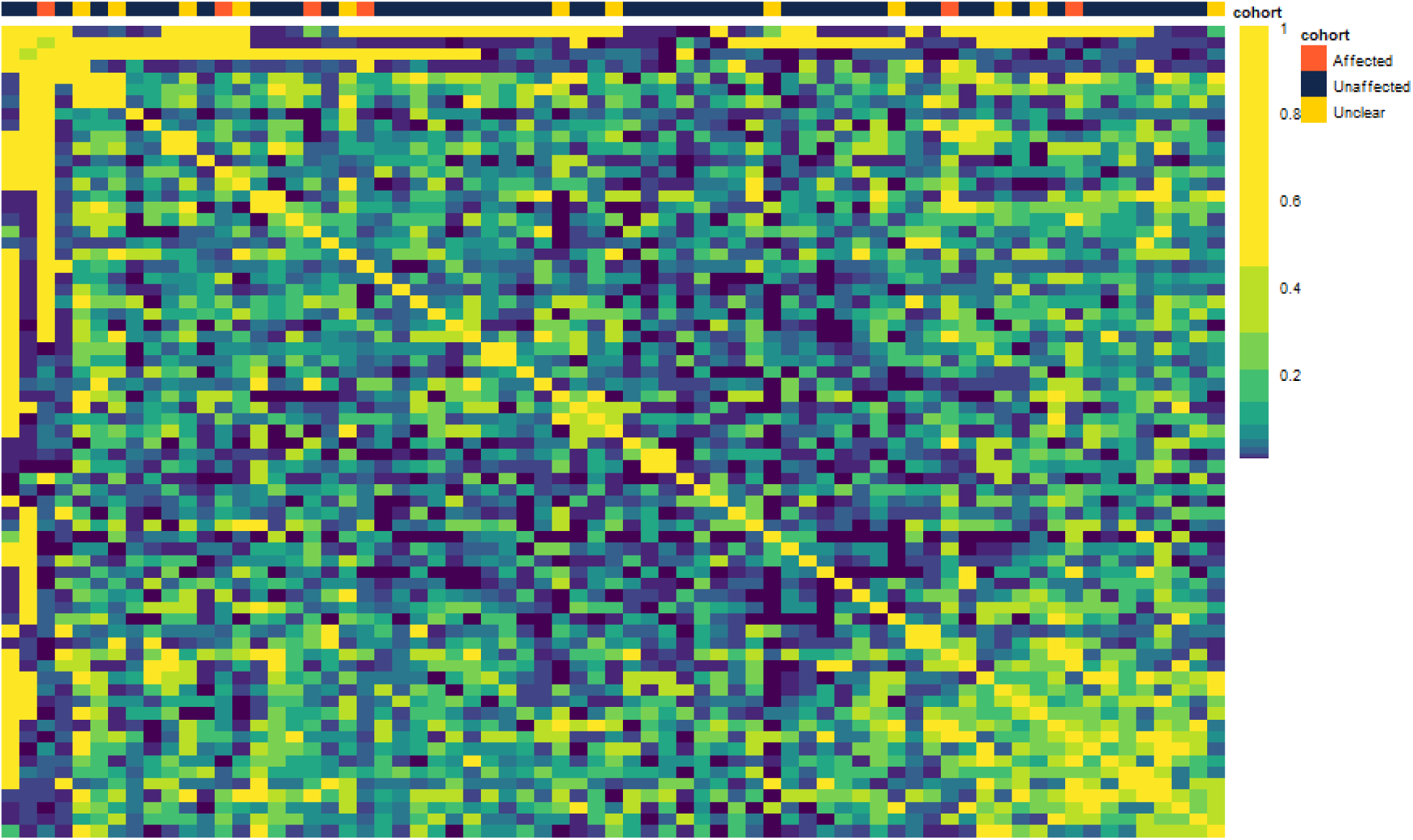
Pairwise comparisons of sequence overlap between participants using the Morisita-Horn index revealed few sequence overlaps. Shown are affected (n=22 LOAD+MCI in orange), unaffected (n=35 in navy), and unclear (n=12 in yellow) as indicated at the bottom of the figure. Each box represents a pairwise Morisita-Horn comparison, which ranges from 0 (little to no sharing of sequences) to 1 (complete sharing).

Given TCRs interact with the MHC complex as part of the adaptive immune response, we imputed and analyzed both class I and II HLA alleles in the Amish participants. After imputation and post-imputation filtering, we observed unique HLA profiles across consensus groups with detailed HLA allele frequencies by each group included in the Supplementary Table 2. Among class I HLA alleles, HLA-A alleles were generally the most frequent, with HLA-A*02:01 representing the highest frequency across all consensus groups. For class II HLA alleles, HLA-DPB1*04:01 had the highest frequency among unaffected, unclear, MCI, and LOAD groups, while HLA-DQB1*03:01 was most frequent in the CINAD group (Supplementary Table 2).

We performed HLA association testing using unadjusted as well as age- and sex-adjusted models, comparing affected (LOAD+MCI) to cognitively unaffected participants. While no HLA allele was significantly associated after multiple testing correction p<3.1×10^-4^), we observed several suggestive associations (Table 4; Figure 2). Among those, HLA-A*03:01 showed marginal significance (p=0.04) in unadjusted analysis but weakened after the covariate adjustment (p=0.06, Figure 2a). HLA-DRB1*15:01 and 06:02, associated with LOAD risk in European ancestry groups (Mansouri et al., 2015), also reached suggestive significance.

**Figure 2.**
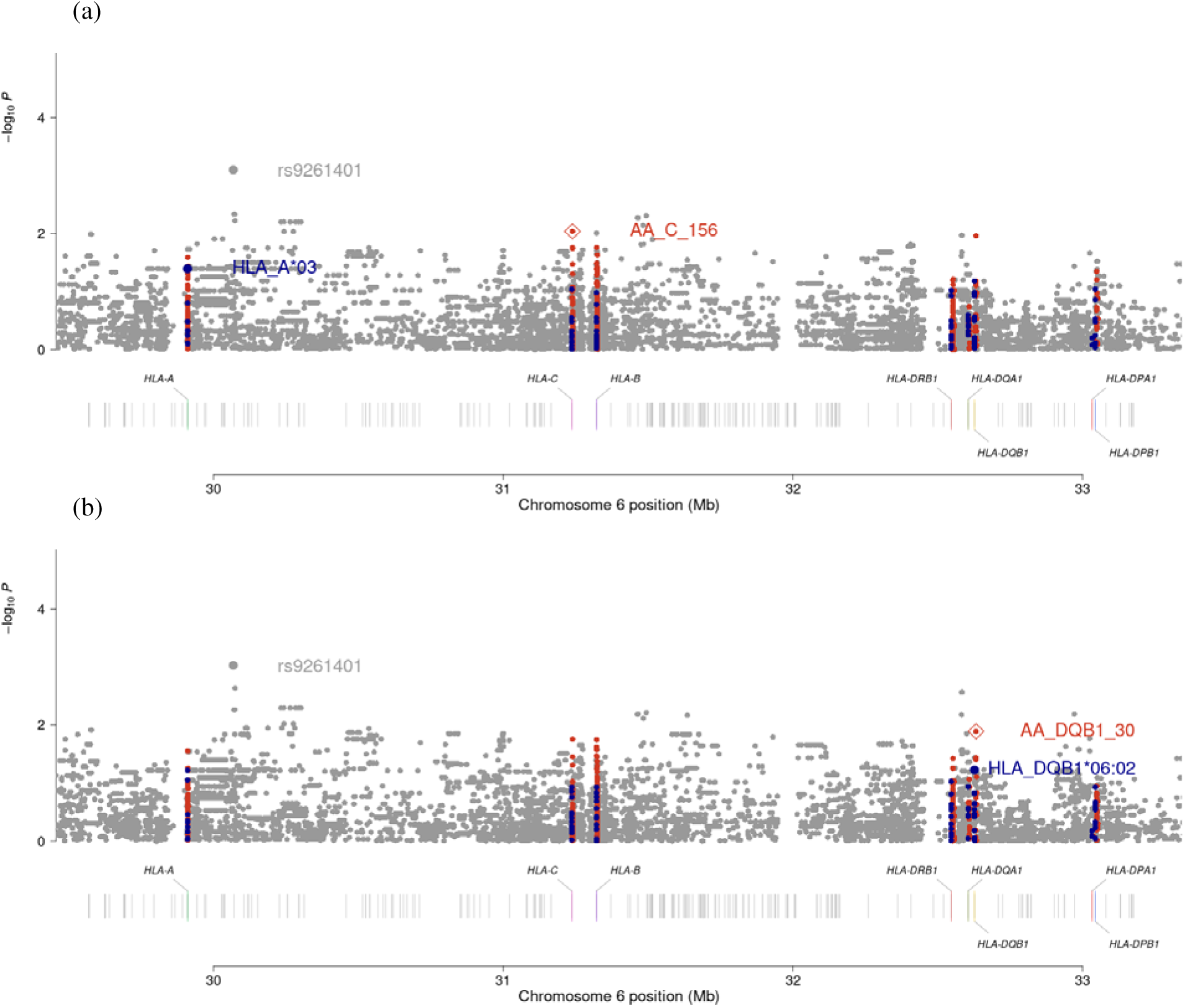
Manhattan plots for HLA tests of association in an Amish aging cohort study. (a) Unadjusted and (b) adjusted (age, sex) association test results comparing affected (LOAD+MCI) to unaffected participants. Points represent individual test results: classical HLA alleles (navy), HLA amino acid variants (orange), and SNPs (grey), plotted by their position on chromosome 6 against their corresponding significance level as shown by -log10(P) on the y-axis. In addition to their chromosome position, the genomic positions of HLA genes are depicted below each Manhattan plot. Only the peak variants within each category are labeled in each association test.

**Table 4.** HLA associations in an Amish aging cohort study. HLA association tests were performed at four-digit resolution, comparing affected (LOAD+MCI; n=22) to unaffected (n=35) participants with results shown for HLA alleles reaching a significance threshold of p<0.1. Both unadjusted **(a)** and adjusted **(b)** (for age at last exam and sex) analyses are presented. Abbreviations: human leukocyte antigen (HLA), odds ratio (OR), p-value (P), and standard error (SE).

We examined potential confounding by autoimmune conditions using participant survey data for type 1 diabetes (T1D), multiple sclerosis (MS), and rheumatoid arthritis (RA). None reported having T1D or MS, and five reported having RA, an autoimmune disease associated with HLA-DRB1 in European-descent populations drawn from the general population. Among the five participants reporting RA, four were classified as unaffected and one as MCI, suggesting that the observed HLA-DRB1 associations are unlikely explained by autoimmune comorbidities.

## Discussion

TCR sequence profiles of 72 Midwestern rural Amish from an ongoing aging cohort study suggest lower sequence diversity among cognitively affected participants compared with unaffected participants and that this lower sequence diversity is associated with higher LOAD plasma biomarker p-tau181 among affected participants. No other patterns, including clonotype sequence similarity, were shared among cognitively affected participants, even in this relatively homogeneous population. Despite the small sample sizes, these data indicate a relationship between the adaptive immune system and LOAD that requires further study to better understand its role in pathogenesis, if any.

A plausible link between the adaptive immune system and dementia is supported by the literature and is related to the phenotypic hallmark of LOAD: the presence and eventual build-up of neurofibrillary tangles and Aβ plaques in the brain (reviewed in (Hampel et al., 2021)). Brain glia and T-cells have been observed in close proximity to plaques in human LOAD cortices (Rogers et al., 1988), and evidence indicates that at some point in the LOAD process, Aβ is deposited into the walls of cerebral blood vessels where T-cells can encounter it (Apatiga-Perez et al., 2022). In a recent study (Gate et al., 2020), characterization of the cerebrospinal fluid T-cell receptor repertoire of patients with mild cognitive impairment and LOAD offered evidence of clonal expansion of sequences known to bind to Epstein Barr virus (EBV) compared with cognitively normal controls (Gate et al., 2020). Moreover, the presence of T-cells was noted in the brain and around Aβ plaques, suggesting a compromised blood-brain barrier in those with neurodegenerative disorders compared with healthy controls.

Possible involvement of T-cells and the adaptive immune response in the development of LOAD is not limited to Aβ plaque deposition. Recent tau-mediated neurodegeneration mouse models demonstrated the presence of brain microglia and T-cells in neuronal regions affected by tauopathy as compared to mice with Aβ (Chen et al., 2023). Further, repertoire analysis demonstrated TCR clonal expansion specific to tauopathy and neurodegeneration as opposed to Aβ (Chen et al., 2023). Microglia, immune cells of the central nervous system (Butovsky & Weiner, 2018), are known to present antigens in neurodegenerative diseases such as MS (Zrzavy et al., 2017). The tauopathy mouse models identified a microglia antigen presentation phenotype and provided evidence for T-cell infiltration and activation mediated by these microglia (Chen et al., 2023). In humans, comparison of cerebrospinal fluid between cognitively normal and cognitively impaired indicates immune dysregulation and infiltration of antigen-specific T-cells in the brain (Piehl et al., 2022).

In this study of Midwestern Amish, lower TCR diversity measured from peripheral tissue is associated with LOAD+MCI; unsurprisingly, age is a strong confounder of this association. No matter the method used, cross-sectional (Y. Wang et al., 2025) and longitudinal (Sun et al., 2022) studies generally demonstration that TCR diversity is lower among older, healthy individuals compared with younger individuals (Chu et al., 2019), consistent with the concept of immunosenescence or the decline of immune system with age (Nikolich-Žugich, 2018). Using the same technology as described here, TCR sequence diversity of pediatric patients from the NEPTUNE study have very low Simpson’s clonality (and diverse TCR repertoires) (Liu et al., 2023). Immunocompromised individuals also have low TCR sequence diversity, and by extension, so do individuals with chronic diseases that affect the immune response (e.g., chronic kidney disease (Crawford et al., 2018)). Neither sex nor *APOE e4* status differed across consensus group phenotypes and consequently were not further explored as modifiers of TCR diversity.

While not associated with affection status independent of age, we found that TCR sequence diversity was associated with p-tau181, a biomarker associated with LOAD pathology (Salami et al., 2022). A recent study of Amish, of which the current study is a subset, also showed elevated p-tau181 levels among the affected individuals as compared to cognitively normal participants, particularly among the *APOE e4* carriers (P. Wang et al., 2025). Our findings further indicate that reduced TCR diversity is associated with higher p-tau181 levels even after adjusting for age and sex among the affected group. In contrast, the unaffected group shows the opposite pattern, with higher p-tau181 associated with decreased clonality (maintained diversity). The opposing results, which are uncorrected for multiple testing, may be due to the small sample sizes tested here. Despite limited statistical power, our results provide evidence that the adaptive immune system has a role in or is a marker of LOAD pathogenesis.

The adaptive immune system response to antigens relies on interaction between TCR and MHC antigen-presenting cells. For the Amish in the present study, the predominant Class I allele across all LOAD consensus diagnosis groups was HLA-A*02:01 (Supplementary Table). This distribution aligns with a prior study that focused on the genetically isolated Dariusleut Hutterites where HLA-A*02 and HLA-B*05/08 were identified as the most frequent based on typed HLA alleles via microlymphocytotoxicity testing (Morgan et al., 1980). Prior research demonstrated that haplotype HLA A*03:01∼B*07:02∼DRB1*15:01∼DQA1*01:02∼DQB1*06:02 conferred strong LOAD risk among participants of European ancestry (Steele et al., 2017), while large-scale genome-wide association studies (GWAS) and meta-analyses have consistently implicated rs9271192, which maps to HLA-DRB1, as a LOAD risk factor (Jansen et al., 2019; Lambert et al., 2013; Lu et al., 2017; Moreno-Grau et al., 2019). Although not statistically significant after adjustment for age and sex or multiple testing, this Amish study highlighted similar alleles: HLA-A*03:01, HLA-DRB1*15:01, and HLA-DQB1*06:02. These associations, however, are in the opposite direction compared with previous studies, suggesting potential differences in HLA LOAD risk in the Midwestern Amish compared with participants of European descent drawn from the general population.

This study has several limitations. The present study was powered only for large differences and effect sizes. TCRs sequenced here are from the periphery and may not represent TCRs in cerebrospinal fluid or the brain. Indeed, studies in MS (de Paula Alves Sousa et al., 2016) and LOAD are unclear to what extent blood TCR repertoires represent cerebrospinal repertoires, for example. Also, only the beta chain was targeted for sequencing. Nevertheless, sequences described here are likely representative of TCR diversity given 1) only one beta chain is expressed on each T-cell (Uematsu et al., 1988), 2) the beta chain is longer and contains greater sequence diversity compared with the alpha chain (Woodsworth et al., 2013), and 3) the beta chain frequently interacts with the antigen (Glanville et al., 2017). Finally, rare sequences such as those observed only once in millions of cells sequenced may not be represented in the final datasets (Robins, 2013; Woodsworth et al., 2013).

Future studies with larger samples sizes and more diverse groups are needed to better understand the role of adaptive immunity in LOAD development. T-cell depletion in tauopathy mice prevents neurodegeneration (Chen et al., 2023), making the adaptive immune system both an attractive therapeutic target as well as an attractive biomarker for early detection of brain atrophy. As incidence and prevalence are projected to increase in the coming decades, the need persists for a better understanding of neurodegenerative disease risk and more precise measures to prevent, treat, and cure it.

## Data Availability

Data produced in the present study are available upon reasonable request to the authors.

## Acknowledgments

We thank Dr. William Bush for helpful comments on this manuscript. This work was supported by 3RF1AG058066, 5R03AG063229, and 5R01AG058066. LAC was supported by T32HL007567, and SL received a scholarship from the Diana Jacobs Kalman/AFAR for Research in the Biology of Aging. This project was also supported by the Clinical and Translational Science Collaborative of Northern Ohio which is funded by the National Center for Advancing Translational Sciences (NCATS) of the National Institutes of Health, UM1TR004528. The content is solely the responsibility of the authors and does not necessarily represent the official views of the NIH.

## Author Contributions

**Jessica N. Cooke Bailey** Supervision, Writing – Review & Editing; **Dana C. Crawford** Conceptualization, Formal Analysis, Investigation, Writing – Original Draft, Supervision, Project Administration, Funding Acquisition; **Lauren A. Cruz** Formal Analysis, Investigation, Visualization, Writing – Review & Editing; **Sarada L. Fuzzell** Resources, Writing – Review & Editing; **Jonathan L. Haines** Resources, Writing – Review & Editing, Project Administration, Funding Acquisition; **Sherri D. Hochstetler** Resources, Writing – Review & Editing; **Tyler Kinzy** Formal Analysis, Investigation, Data Curation, Visualization, Writing – Review & Editing; **Ioanna Konidari** Resources, Writing – Review & Editing; **Renee A. Laux** Resources, Writing – Review & Editing; **Alan J. Lerner** Resources, Writing – Review & Editing; **Shiying Liu** Formal Analysis, Investigation, Data Curation, Visualization, Writing – Review & Editing; **Audrey Lynn** Resources, Supervision, Writing – Review & Editing; **Jacob L. McCauley** Resources, Supervision, Writing – Review & Editing; **Penelope Miron** Resources, Writing – Review & Editing; **Kristy L. Miskimen** Resources, Writing – Review & Editing; **Paula K. Ogrocki** Resources, Writing – Review & Editing; **Margaret A. Pericak-Vance** Resources, Writing – Review & Editing, Project Administration, Funding Acquisition; **William K. Scott** Resources, Writing – Review & Editing, Project Administration, Funding Acquisition; **Yeunjoo E. Song** Data Curation, Resources, Writing – Review & Editing

---

**Supplementary Table 1.** Summary of sequence-level metrics, by consensus phenotype status. All metrics save for Shannon diversity were provided by Adaptive Biotechnologies ANALYZER bioinformatic pipeline. Shown are means (standard deviations). Unaffected participants were compared with affected participants (LOAD+MCI) using Welch’s t-test. Abbreviations: Late-onset

**a)**
| **Unadjusted** |  |  |
| --- | --- | --- |
| **HLA** | **OR (SE)** | **P** |
| A*03:01 | 0.25 (0.68) | 0.0402 |
| DQB1*06:02 | 0.23 (0.81) | 0.0658 |
| DPB1*04:01 | 0.48 (0.43) | 0.0906 |
| C*04:01 | 2.84 (0.62) | 0.0911 |
| DRB1*15:01 | 0.25 (0.83) | 0.0951 |

**b)**
| **Adjusted** |  |  |
| --- | --- | --- |
| **HLA** | **OR (SE)** | **P** |
| DQB1*06:02 | 0.20<br>(0.86) | 0.0594 |
| A*03:01 | 0.28<br>(0.68) | 0.0606 |
| A*33:03 | 8.52<br>(1.26) | 0.0885 |
| DRB1*15:01 | 0.23<br>(0.88) | 0.0941 |

**Supplementary Table 2.**
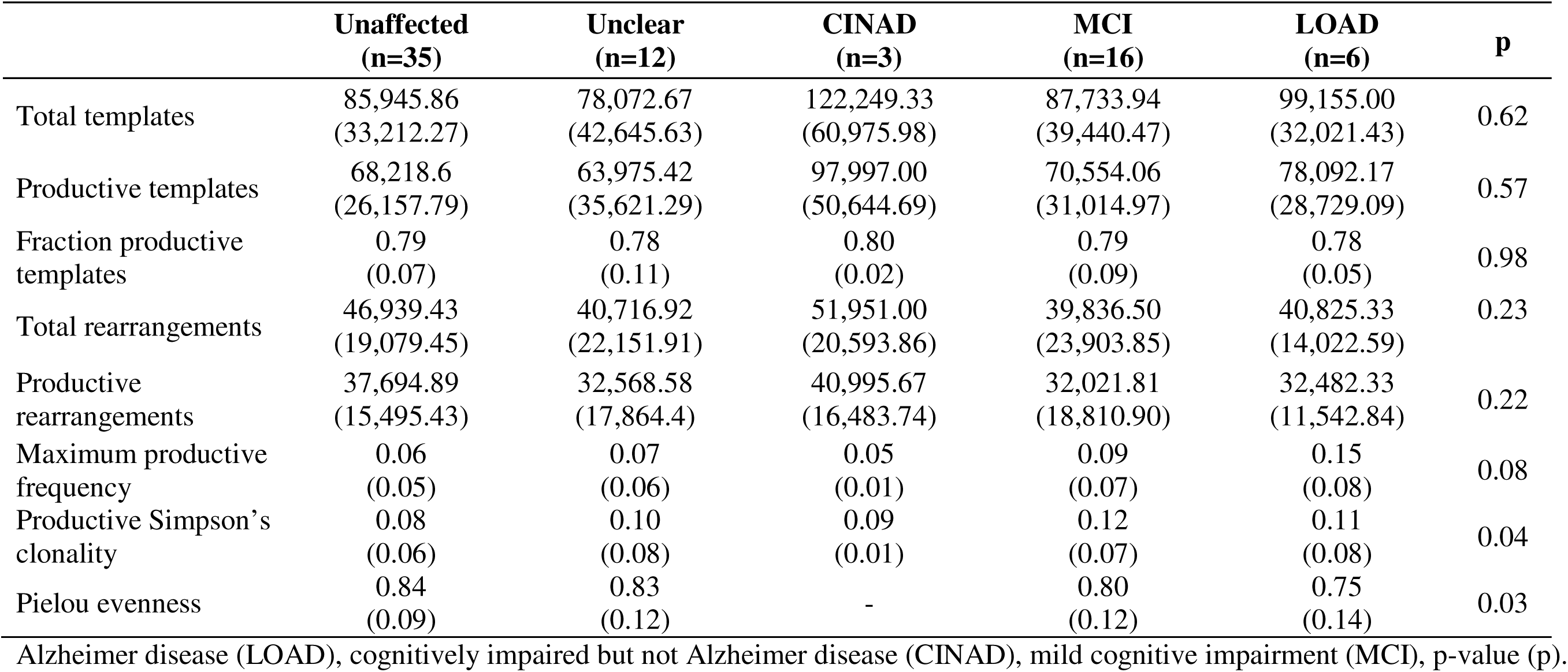
HLA allele frequency, by consensus phenotype status in an Amish aging cohort study. HLA at four-digit resolution. Abbreviations: Late-onset Alzheimer disease (LOAD), cognitively impaired but not Alzheimer disease (CINAD), mild cognitive impairment (MCI).

|  | **Unaffected**<br>**(n=35)** | **Unclear**<br>**(n=12)** | **CINAD**<br>**(n=3)** | **MCI**<br>**(n=16)** | **LOAD**<br>**(n=6)** | **p** |
| --- | --- | --- | --- | --- | --- | --- |
| Total templates | 85,945.86<br>(33,212.27) | 78,072.67<br>(42,645.63) | 122,249.33<br>(60,975.98) | 87,733.94<br>(39,440.47) | 99,155.00<br>(32,021.43) | 0.62 |
| Productive templates | 68,218.6<br>(26,157.79) | 63,975.42<br>(35,621.29) | 97,997.00<br>(50,644.69) | 70,554.06<br>(31,014.97) | 78,092.17<br>(28,729.09) | 0.57 |
| Fraction productive<br>templates | 0.79<br>(0.07) | 0.78<br>(0.11) | 0.80<br>(0.02) | 0.79<br>(0.09) | 0.78<br>(0.05) | 0.98 |
| Total rearrangements | 46,939.43<br>(19,079.45) | 40,716.92<br>(22,151.91) | 51,951.00<br>(20,593.86) | 39,836.50<br>(23,903.85) | 40,825.33<br>(14,022.59) | 0.23 |
| Productive<br>rearrangements | 37,694.89<br>(15,495.43) | 32,568.58<br>(17,864.4) | 40,995.67<br>(16,483.74) | 32,021.81<br>(18,810.90) | 32,482.33<br>(11,542.84) | 0.22 |
| Maximum productive<br>frequency | 0.06<br>(0.05) | 0.07<br>(0.06) | 0.05<br>(0.01) | 0.09<br>(0.07) | 0.15<br>(0.08) | 0.08 |
| Productive Simpson's<br>clonality | 0.08<br>(0.06) | 0.10<br>(0.08) | 0.09<br>(0.01) | 0.12<br>(0.07) | 0.11<br>(0.08) | 0.04 |
| Pielou evenness | 0.84<br>(0.09) | 0.83<br>(0.12) | - | 0.80<br>(0.12) | 0.75<br>(0.14) | 0.03 |

